# Childhood Stunting and Its Associated Factors in Ethiopia

**DOI:** 10.1101/2022.05.03.22274629

**Authors:** Woldemariam Erkalo Gobena

## Abstract

Child malnutrition is the root cause of nearly half (45%) of all child deaths, especially in low-income nations. Ethiopia has the highest frequency of stunting among Sub-Saharan African countries, at 38 percent. The study’s major goal was to use cluster specific models to identify risk factors for stunting in Ethiopian children under the age of five. The data was gathered from the EDHS 2016, a nationally representative survey of children aged 0 to 59 months. The research was carried out using generalized linear mixed models from the cluster specific model family. The variables child’s age, mother’s education level, mother’s BMI, place of residence, wealth index, and prior birth interval were determined to be important drivers of child malnutrition in Ethiopia as a result of the analysis. According to the findings, children with a shorter previous birth interval (less than 24 months) were more likely to be stunted than those with a longer previous birth interval. Rural Ethiopian children were more likely than urban Ethiopian children to be stunted. It is advised that in order to reduce childhood malnutrition, parents’ awareness and implementation of proper young child feeding practices, as well as frequent growth monitoring and appropriate and timely interventions, should be prioritized.

## INTRODUCTION

Malnutrition in children is caused by a lack of food, diarrhea and other diseases, poor sanitation, and a lack of parental education (1, 2). Food insecurity, inadequate mother and child care, and poor health services and the environment all contribute to poor diets and disease (3). These elements have a measurable negative impact on body function and clinical outcome (4). The majority of anthropometric deficiencies in children under the age of five are caused by this problem in the world’s least developed countries (5).

Despite known solutions, child malnutrition remains a major global public health issue (6). Child malnutrition is the underlying cause of nearly half of all child deaths (45%) in developing nations, particularly in poor socioeconomic communities (7). In 2018, an estimated 149 million children under the age of five were stunted globally, with another 49 million wasting (8).

Stunting is on the decline in Sub-Saharan Africa, yet it still affects over 30% of the population (8). Several African countries have changed their national nutrition policies, strategies, and action plans in order to combat malnutrition (9). Ethiopia has the highest rate of wasting in Sub-Saharan Africa, at 10%, and the highest rate of stunting at 38%. There is regional variation in stunting within Ethiopia. Child stunting is most prevalent in Amhara, Benishangul-Gumuz, Afar, and Dire Dawa. Drought-related public health and nutrition difficulties were reported in hard-to-reach districts of Afar, Somalia, Benishangul-Gumuz, and Gambela (8). The present rate of improvement is insufficient to meet the World Health Organization’s (WHO) global goal of a 40% reduction in stunted children by 2025 (10).

The goal of this study is to use cluster specific models to identify risk factors for stunting in Ethiopian children under the age of five.

## MATERIALS AND METHODS

### Data Source

The Ethiopian Demographic and Health Survey 2016 provided information on childhood stunting. It was gathered by Ethiopia’s Central Statistical Agency between January 18 and June 27, 2016.

### Study Population and Variables

A total of 10, 641 children under the age of 59 months were found in the households of selected clusters for this study. A total of 4,621 children and 626 clusters had their complete height-for-age records collected. On the remaining 6,020 children and 19 clusters’ height-for-age records, there were missing values. Stunting is employed as a response variable in this study, and it is measured using the height-for-age Z-score (multiplied by 100). Thus, children whose height-for-age Z-score is less than minus two standard deviations (−2 SD) from the median are termed stunted, whereas children whose height-for-age Z-score is greater than minus two standard deviations (−2 SD) from the median are considered not-stunted. (5).

The following fourteen factors were used as proxy indicators of socioeconomic, demographic, health, and environmental characteristics: mother’s education level, wealth index, number of household members, place of residence, child’s age, child’s sex, mother’s age at child’s birth, number of antenatal care visits, previous birth interval, birth order, mother’s BMI, diarrhea, fever, and source of drinking water.

### Methods of Data Analysis

The response variables in a generalized linear model (GLM) are assumed to be independent. However, in clustered data, observations are typically obtained from the same unit, resulting in a cluster of correlated observations. In the EDHS, for example, the dependent variable (stunting) was measured once for each region’s representative samples nested within clusters. As a result, GLM has been extended to cluster specific models for this purpose.

### Cluster (Subject) Specific Models

Parameters relevant to clusters or persons within a population are included in cluster specific models. As a result, random effects are used directly in modeling random variation in the dependent variable at various levels of the data.

### Generalized Linear Mixed Model (GLMM)

The generalized linear mixed model (GLMM) is a type of cluster specific model that adds non-normal responses and a link function to ordinary regression. The extended linear mixed model is an expansion of the linear predictor that allows for both random and fixed effects (11).

Let *y*_*ij*_ denote the response of *i*^*th*^individual child from j^th^ cluster where *i* = 1,2,…, *n*_*j*_ *and Y*_*j*_ *is the n*_*j*_ dimensional vector of all measurements available for cluster *j*. Let *f*(*b_j_/D*) be the density of the *N*(0, *D*) distribution for the random effects *b*_*j*_. Assumed conditionally on q-dimensional random effects *b*_*j*_ to be drawn independently from *N*(0, *D*), the outcomes *y*_*ij*_ of *Y*_*j*_ are independent with the density of the form

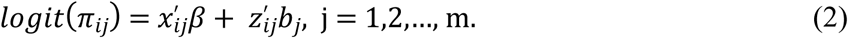

Then the generalized linear mixed model (12); with logit link is defined as

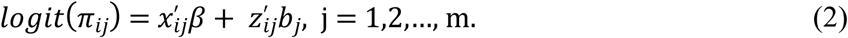

Where, *E*(*Y*_*ij*_|*b*_*j*_) = *π*_*ij*_, is the mean response vector conditional on the random effects *b*_*j*_, for children in cluster j and, *X*_*ij*_ *and Z*_*ij*_ are *p*-dimensional and *q*-dimensional vectors of known covariate values. The random effects *b*_*j*_ are assumed to follow a multivariate normal distribution with mean *0* and covariance matrix *D*.

## RESULTS

### Descriptive Statistics

Of the 4,621 children who were included in this study, 50.9% were male and 49.10% were female children. Out of these children, 36.01% male children and 33.76% female children were stunted. Table 1 shows the descriptive results.

**Table 1:**
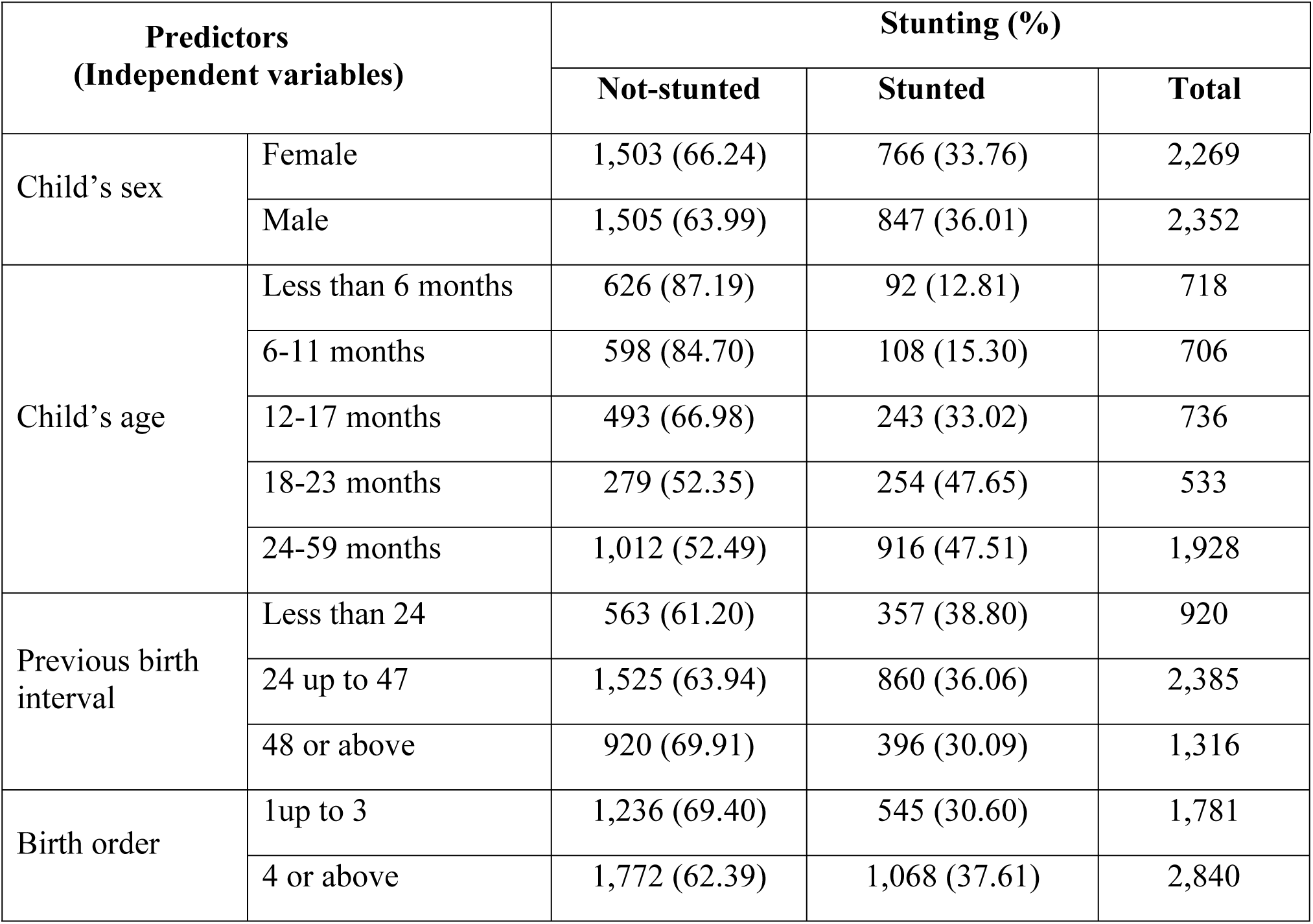
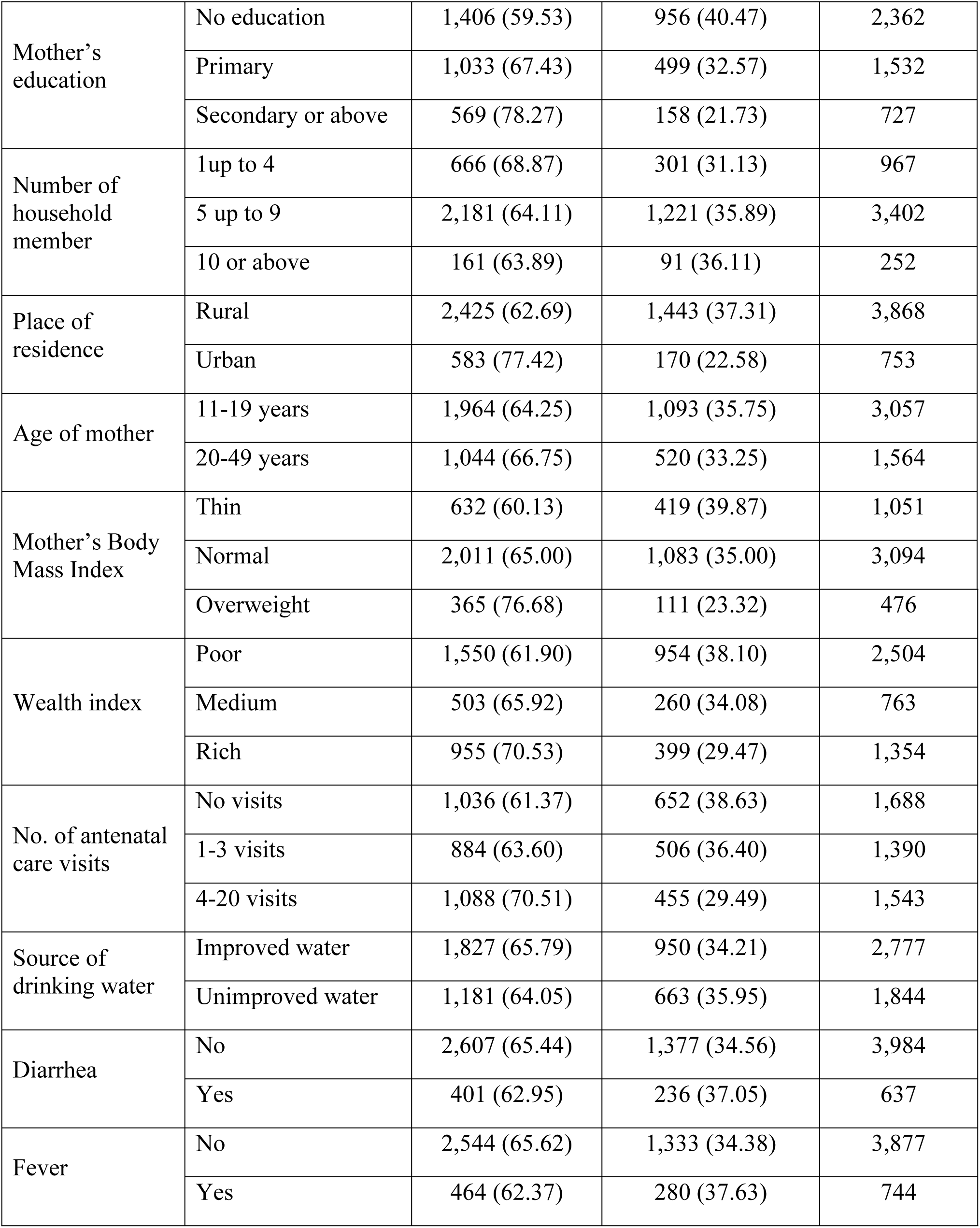
Cross Tabulation of Prevalence of Stunting by Maternal, Socioeconomic and Demographic Variables.

**Table 2:** Information Criteria for Comparison of One and Two Random Intercept Models.

| Models | AIC | BIC | LogLik | Deviance | $\sigma_W$ | $\sigma_B$ | P |
| --- | --- | --- | --- | --- | --- | --- | --- |
| **Model with one random intercept** | 5325.9 | 5422.4 | -2647.9 | 5295.9 | 0.5088 |  |  |
| **Model with two random intercepts** | 5314.8 | 5413.8 | -2639.9 | 5279.8 | 0.4398 | 0.2366 | <.0001 |

### Analysis of Generalized Linear Mixed Model (GLMM)

#### Model Building in GLMM

Model fitting in the GLMM begins with the acceptance of marginal model covariates. The model also includes random effects, in this example, random intercepts, to account for differences across and within regions. First, the main effect covariates and the two random intercepts models were fitted and non-significant covariates were removed sequentially, starting with the variables with the greatest p-value for fixed effect covariates, as is customary. The saturated GLMM models were fit as follows.

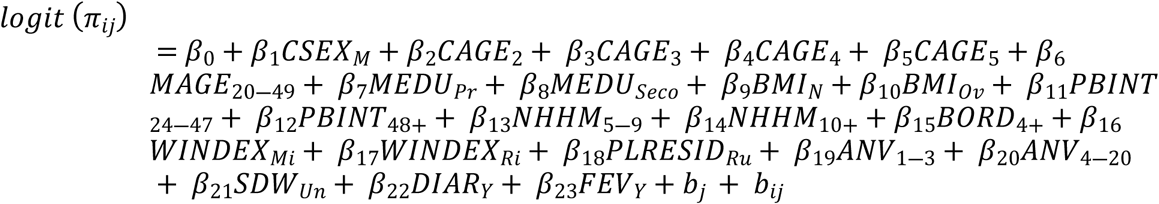

Where, *b*_*j*_ and *b*_*ij*_ are the two random intercepts.

Two models were fitted to determine which of the two random effects models was better: one with two random intercepts to estimate between and within regional variations, and the other with one random intercept model to estimate within regional variation. The AIC and the Likelihood ratio test (LRT) were used to compare the two models and choose the best one.

Where, σ_W_ and σ_B_ are within and between regional standard deviation respectively. The AIC of the model with two random intercepts is lowered from 5325.9 to 5314.8, and the deviance is reduced from 5295.9 to 5279.8, as shown in table 3. The log likelihood ratio test’s low p-value (P< 0.0001) further implies that the model with two random intercepts is parsimonious. The p-value of the two models’ log likelihood ratio test is P. When a model without random effects is studied (i.e. just the generalized linear model), the AIC value is 5354.5, which is large when compared to the two models with random effects above.

**Table 3:**
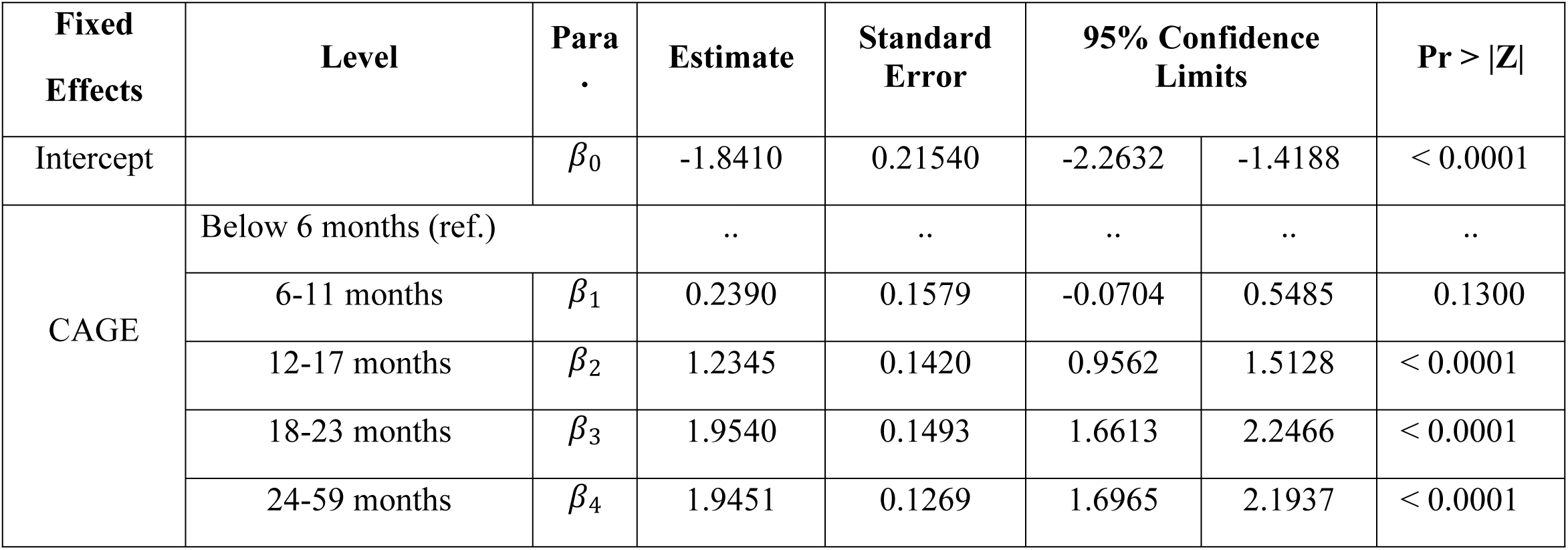
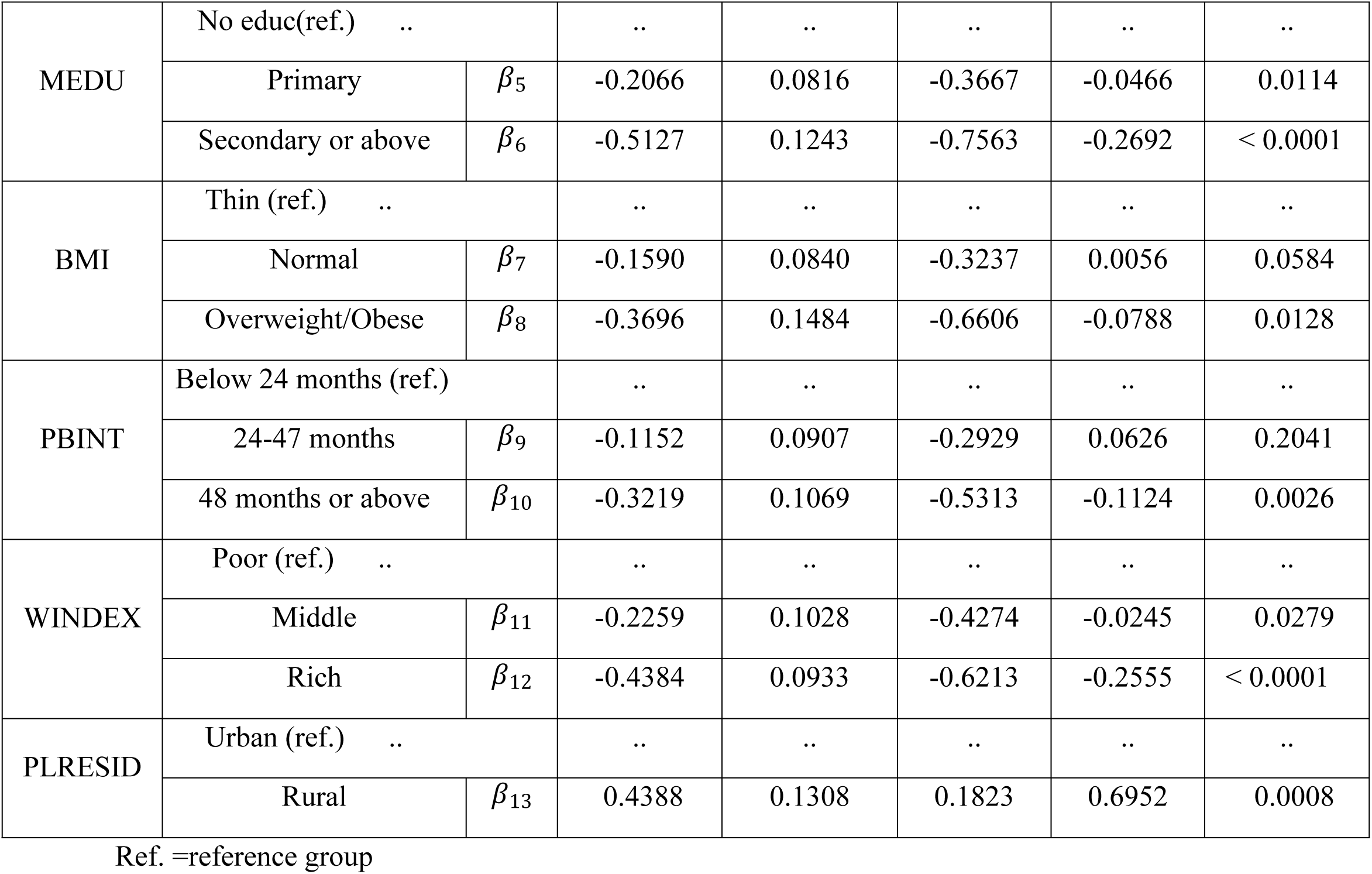
Parameter Estimates (Standard Errors) and Corresponding P-Value for GLMM.

| Fixed Effects | Level | Para . | Estimate | Standard Error | 95% Confidence Limits |  | Pr > \|Z\| |
| --- | --- | --- | --- | --- | --- | --- | --- |
| Intercept | | $\beta_0$ | -1.8410 | 0.21540 | -2.2632 | -1.4188 | < 0.0001 |
| CAGE | Below 6 months (ref.) |  | .. | .. | .. | .. | .. |
| | 6-11 months | $\beta_1$ | 0.2390 | 0.1579 | -0.0704 | 0.5485 | 0.1300 |
| | 12-17 months | $\beta_2$ | 1.2345 | 0.1420 | 0.9562 | 1.5128 | < 0.0001 |
| | 18-23 months | $\beta_3$ | 1.9540 | 0.1493 | 1.6613 | 2.2466 | < 0.0001 |
| | 24-59 months | $\beta_4$ | 1.9451 | 0.1269 | 1.6965 | 2.1937 | < 0.0001 |
| MEDU | No educ(ref.) .. |  | .. | .. | .. | .. | .. |
| | Primary | $\beta_5$ | -0.2066 | 0.0816 | -0.3667 | -0.0466 | 0.0114 |
| | Secondary or above | $\beta_6$ | -0.5127 | 0.1243 | -0.7563 | -0.2692 | < 0.0001 |
| BMI | Thin (ref.) .. |  | .. | .. | .. | .. | .. |
| | Normal | $\beta_7$ | -0.1590 | 0.0840 | -0.3237 | 0.0056 | 0.0584 |
| | Overweight/Obese | $\beta_8$ | -0.3696 | 0.1484 | -0.6606 | -0.0788 | 0.0128 |
| PBINT | Below 24 months (ref.) |  | .. | .. | .. | .. | .. |
| | 24-47 months | $\beta_9$ | -0.1152 | 0.0907 | -0.2929 | 0.0626 | 0.2041 |
| | 48 months or above | $\beta_{10}$ | -0.3219 | 0.1069 | -0.5313 | -0.1124 | 0.0026 |
| WINDEX | Poor (ref.) .. |  | .. | .. | .. | .. | .. |
| | Middle | $\beta_{11}$ | -0.2259 | 0.1028 | -0.4274 | -0.0245 | 0.0279 |
| | Rich | $\beta_{12}$ | -0.4384 | 0.0933 | -0.6213 | -0.2555 | < 0.0001 |
| PLRESID | Urban (ref.) .. |  | .. | .. | .. | .. | .. |
| | Rural | $\beta_{13}$ | 0.4388 | 0.1308 | 0.1823 | 0.6952 | 0.0008 |

Ref. =reference group

Following that, the variables for the fixed effect were evaluated, and potential covariates were chosen by gradually deleting covariates with the greatest p-value. In addition, a model with a small number of covariates is seen to be better. As a result, the final GLMM for childhood stunting is as follows:

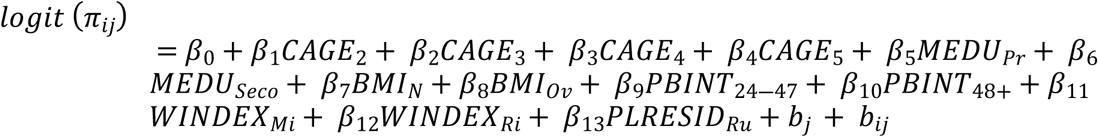

### Parameter Interpretation of GLMM

Parameter interpretation in GLMM analysis is dependent on specific subjects or clusters. The random effects, which are common for all individual children in the same cluster, are used to interpret the parameters.

When compared to children from illiterate mothers, children from mothers who have learnt primary education and secondary education or higher were exp(−0.2066)= 0.8133 (95 percent CI: 0.6931, 0.9545) and exp(−0.5127)= 0.5989 (95 percent CI: 0.4694, 0.7640) times less likely to be stunted, respectively. When compared to children from uneducated mothers in the same cluster, childhood stunting is reduced by 18.67 percent and 40.11 percent, respectively, for children whose mothers attended primary and secondary education or higher.

Children with mothers who were overweight (BMI ≥ 25.0) were less likely to be stunted. Children of overweight mothers had a 31% lower risk of being stunted than children of thin mothers in the same cluster, according to the estimated odds ratio (OR=0.6910; 95 percent CI: 0.5166, 0.9243).

Children with a birth interval of 48 months or more had a 27.52 percent lower chance of being stunted than children with a birth interval of less than 24 months in the same cluster, according to the calculated odds ratio (OR=0.7248; 95 percent CI: 0.5878 0.8937).

Children from rural residences were also 55.08 percent (OR=1.5508, 95 percent CI: 1.2000, 2.0042) more likely to be stunted than children from urban residences at the given constant random effect.

## Discussion

Based on EDHS 2016 data, the goal of this study was to identify the risk factors for stunting in Ethiopian children aged five and under. The height-for-age z scores were used to assess stunting. A variety of summary statistics were used as a preliminary analysis to investigate the relationship between the response variable of interest and the available covariates. It should be noted that the conclusions drawn from the analysis of various summary statistics are inconsistent, which could be related to the fact that they employ different amounts of data, which affects the power of their findings. As a result, various statistical approaches were used to account for the clustered character of correlated observations in the analysis. After that, the data was examined using a random effects model.

The goal of GLMM was to assess differences in stunting within and between Ethiopian regions. Two models were fitted; one with just one random intercept to examine only within regional variation and the other with two random intercepts to account for both within and between regional variations. The AIC values were used to compare the two models, followed by a likelihood ratio test, and the model with two random intercepts was found to be more favorable. This illustrates the importance of taking into account both within and between regional variations when analyzing stunting, as well as the existence of within and between regional heterogeneity in children stunting. This outcome is in line with the explanation of (13).

Age of child, previous birth interval, mother’s education level, place of residence, wealth index, and mother’s BMI were all found to be significantly associated with stunting, whereas sex of child, age of mother at first birth, number of household members, number of antenatal care visits, birth order, source of drinking water, had diarrhea recently, and had fever in the two weeks prior to survey were not.

The results of this study revealed that the risk of stunting rose as children grew older. When compared to children under the age of six months, children aged 18-23 months had a considerably higher risk of stunting. Ethiopia, Bangladesh, Madagascar, and Malawi have all published similar studies (14-18). It could be related to the incorrect and late introduction of low nutritional quality supplementary food (19), and a big percentage of guardians in rural areas are ignoring their children’s optimal food requirements as they get older (20). This finding is also in line with the findings of the EDHS 2016 national report, which found that the prevalence of stunting rises as a child’s age rises (21).

The prevalence of stunting has a significantly substantial and inverse association with the previous birth gap. Children who had a longer interval between births had a lower probability of becoming stunted. Both birth interval groups (24 to 47 months and 48 months or more) are statistically significant, according to the results of this study. As a result, the likelihood of becoming stunted diminishes as the birth interval grows. The majority of past research backs up this conclusion (22-25). The large and increased risk of stunting among infants born at shorter intervals could be attributed to women’s nutritional resources being depleted during pregnancy and lactation (26). If the woman falls pregnant too soon again, close spacing may have a negative impact on the prior kid, who may be prematurely weaned. Rural children were shown to be the most affected by stunting in this study. This could be related to the narrow spacing and low prevalence of contraception in rural areas. This contradicts the findings of a research done by (17, 27). Using multivariable multilevel logistic regression and ordinary logistic regression, they found no significant relationship between birth order and stunting for the entire dataset.

The mother’s BMI, which is calculated by dividing her weight in kilograms by the square of her height in meters, was found to be adversely associated with childhood stunting in this study. Thin mothers (BMI <18.5) are malnourished themselves, and their children are more likely to be stunted. A number of researches have come to the same conclusion. On average, mothers with a low BMI have kids with a low birth weight (17, 28, 29). This finding is also consistent with certain African research (30, 31), which show that a mother’s nutritional health influences her ability to effectively carry, birth, and care for her children. Women who are undernourished (thinness or obesity) may have trouble giving birth and may give birth to malnourished children. The findings suggest that a link exists between the mother’s thinness and the child’s nutritional status.

Childhood stunting was discovered to be inversely connected to the mother’s educational level. Children whose moms have a secondary or higher level of education have a much lower risk of stunting than children whose mothers have no education status. This finding seems to be in line with the findings of other investigations (17, 32, 33). These findings highlight the relevance of girls’ education as an alternative technique for combating childhood stunting and promoting healthy eating habits for early children. Increased awareness of sanitation methods and healthy habits, as well as higher levels of maternal education, can help to minimize childhood stunting in various ways.

Childhood stunting was also discovered to be inversely connected to the home wealth index. Children from poor families are reported to be at a higher risk of stunting than children from wealthy families (14, 34-36).

According to the findings, urban children are less likely to be stunted (malnourished) than their rural counterparts because the quality of the health environment and sanitation is better in cities, whereas living conditions in rural areas are associated with poor health and a lack of personal hygiene, both of which are risk factors for malnutrition. This is in line with several research, which show that the mother’s place of residence has a statistically significant impact on the nutritional health of her children (17, 37).

## CONCLUSIONS

The goal of this study was to use cluster specific models to identify risk variables for children stunting using a dataset of childhood stunting obtained from the Central Statistics Agency. As a result, there is a wide range of childhood stunting between and within regions. The study found that child’s age, mother’s educational status, mother’s BMI, preceding birth interval, wealth status, and place of residence all had a significant impact on childhood stunting, whereas child’s sex, mother’s age at first birth, number of household members, number of antenatal care visits, birth order, source of drinking water, diarrhea recently, and fever in the two weeks prior to survey were not. More importantly, this research adds to our knowledge of the individual and collective effects of maternal, socioeconomic, and child-related risk factors in Ethiopian childhood stunting.

## Data Availability

The data is available at URL: https://dhsprogram.com

## Abbreviations

AIC: Akaike’s Criterion
CSA: Central Statistical Agency
EDHS: Ethiopian Demographic and Health Survey
UNICEF: United Nations Children’s Fund
SD: Standard Deviation

## Acknowledgements

I acknowledge the Ethiopian Central Statistical Agency (CSA) for providing me the data. I also gratefully acknowledge all authors listed in the references.

